# Molecular epidemiology of human rhinovirus from one-year surveillance within a school setting in rural coastal Kenya

**DOI:** 10.1101/2020.03.09.20033019

**Authors:** Martha M. Luka, Everlyn Kamau, Irene Adema, Patrick K. Munywoki, Grieven P. Otieno, Elijah Gicheru, Alex Gichuki, Nelson Kibinge, Charles N. Agoti, D. James Nokes

## Abstract

**Background:** Human rhinovirus (HRV) is the most common cause of the common cold but may also lead to more severe respiratory illness in vulnerable populations. The epidemiology and genetic diversity of HRV within a school setting have not been described.

**Objective:** To characterise HRV molecular epidemiology among children attending primary school in a rural location of Kenya.

**Methods:** Between May 2017 to April 2018, over three school terms, we collected 1859 nasopharyngeal swabs (NPS) from pupils and teachers with symptoms of acute respiratory infection in a public primary school in Kilifi County, coastal Kenya. The samples were tested for HRV using real-time RT-PCR. HRV positive samples were sequenced in the VP4/VP2 coding region for species and genotype classification.

**Results:** A total of 307 NPS (16.4%) from 164 individuals were HRV positive, and 253 (82.4%) were successfully sequenced. The proportion of HRV in the lower primary classes was higher (19.8%) than upper primary classes (12.2%), *p*-value <0.001. HRV-A was the most common species (134/253, 53.0%), followed by HRV-C (73/253, 28.9%) and HRV-B (46/253, 18.2%). Phylogenetic analysis identified 47 HRV genotypes. The most common genotypes were A2 and B70. Numerous (up to 22 in one school term) genotypes circulated simultaneously, there was no individual re-infection with the same genotype, and no genotype was detected in all three school terms.

**Conclusion:** HRV was frequently detected among school-going children with mild ARI symptoms, and particularly in the younger age groups (<5-year-olds). Multiple HRV introductions were observed characterised by the considerable genotype diversity.

**Summary points:** We describe the molecular epidemiology of human rhinovirus (HRV) within a school setting over one-year in rural coastal Kenya. A high diversity of HRV infections was observed across all classes with evidence of introduction and transmission of 47 different genotypes.

## INTRODUCTION

Human rhinovirus (HRV) is a frequently detected viral respiratory pathogen ^1^ associated with the common cold ^2,3^, lower respiratory tract infections ^4^ and asthma development and exacerbation ^5,6^. Although the majority of HRV cases are mild and self-limiting, they contribute to substantial economic losses through missed school and workdays ^7,8^. HRV is a common reason for prescribing antibiotics ^9^, potentially contributing to antibiotic resistance. The virus is transmitted via inhalation of contaminated aerosols and close contact with infected persons or surfaces ^10^, and prolonged periods of close contact increase the transmission efficiency ^11^. Children, the elderly and those with pre-existing respiratory conditions have the highest HRV burden in the community ^12–14^.

HRV is a positive sense, single-stranded RNA virus, classified under the genus *Enterovirus* (family *Picornaviridae)* with a genome approximately 7.2 kb long. It is characterised by high genetic diversity and an associated high antigenic diversity ^15^, frustrating vaccine development efforts. There are 169 HRV genotypes distributed across three species: 80 HRV-A genotypes, 32 HRV-B genotypes and 57 HRV-C genotypes ^16^. HRV is non-enveloped and has a spherical capsid with a diameter of about 30 nm which constitutes of 60 copies of each of the four viral proteins: VP4, VP2, VP3 and VP1 ^17^, in that order. Serotype assignment was initially done using monoclonal antibodies by virus neutralisation assays for species A and B ^18,19^. However, HRV-C, discovered in 2006 ^20^, cannot propagate in conventional cell lines limiting serotyping as a classification method ^21,22^. Molecular classification is based on the nucleotide sequence of either the 5’ noncoding region, VP1 or the VP4/2 genome region ^22–24^.

Children constitute a significant reservoir of HRV ^25^. In sub-Saharan Africa, little has been done to investigate the patterns and mechanisms of transmission of HRV in schools and to understand the extent to which school settings contribute to transmission in the community. This is despite that primary school children have the highest contact rates compared to other age-groups in the community ^26^ and play a significant role in infection transmission to their younger siblings ^27,28^. Design of effective intervention strategies against HRV relies on our knowledge of transmission dynamics of the virus in different social networks and population structures ^8,29^. No study has yet explored the diversity and dynamics of HRV infections among school children. This has limited our understanding of the transmission patterns of HRV in this social grouping and the community as a whole.

This study investigated HRV infections in a school setting in rural coastal Kenya by sequence analysis of the VP4/2 junction to describe the frequency, diversity and temporal occurrence of HRV.

## METHODS

### Study area and design

An epidemiological surveillance study was conducted in a rural school located within the Kilifi Health and Demographic Surveillance System (KHDSS) in Kenya ^30^ to characterise the occurrence of respiratory viruses. The study design is described in detail elsewhere^31^. Briefly, the school offers both early childhood development education and primary school education. Pupils and teachers from all classes were enrolled in the study, which took place between May 2017 and April 2018. Pupils were divided into two main groups: the lower primary comprising of day care, kindergarten (KG) levels 1 to 3 and grade one (*n*=5 classes, age range 3-12 years); and the upper primary comprising grades two to eight (*n*=7 classes, age range 7-20 years).

Nasopharyngeal swabs (NPS) were collected from individuals who had at least one of the following acute respiratory illness (ARI) symptoms: cough, sore throat or runny nose. A maximum of 8 samples per class was collected from the lower primary group per week, while a maximum of 4 samples per class was collected from the upper primary group per week. A maximum of 3 samples was collected from the teachers per week. We collected more samples from the lower primary due to the perceived critical role of this age-groups in childhood infectious diseases and hence the need to reduce the level of uncertainty in the estimated risk in this age-groups. Samples were collected in viral transport media (VTM) and transported in cool boxes to the KEMRI-Wellcome Trust Research Programme laboratory where they were stored at -80°C prior to screening. Sampling was suspended during school holidays: months of August, November and December 2017 and from 6^th^ April 2018, which marked the end of the study.

An informed written parental consent for children under the age of eighteen years or individual consent for adults was obtained prior to sample collection. In addition, children whose parents consented were asked for individual assent to participate. Ethical approval was provided by the KEMRI-Scientific Ethics Review Unit (KEMRI-SERU #3332) and the University of Warwick Biomedical and Scientific Research Ethics Committee (BSREC #REGO_2016-1858).

### RNA extraction and rRT-PCR

RNA was extracted from 140 µl of the collected sample using QIAamp® 96 Virus QIAcube HT kit (Qiagen, United Kingdom), according to the manufacturer’s instructions. All samples were tested for HRV using an in-house multiplexed real-time reverse-transcription PCR (rRT-PCR) with a QuantiFast® Multiplex RT-PCR kit (Qiagen, United Kingdom) ^32–34^. A sample was considered HRV positive if the rRT-PCR cycle threshold (*Ct*) value was <35.

### VP4/2 amplification and sequencing

VP4/2 sequencing was used to assign species and genotypes. A genomic region approximately 549 nucleotides and consisting of a hypervariable region of the 5’UTR, the complete VP4 and partial VP2 gene region was amplified for all HRV positive samples using a One-Step RT-PCR kit (QIAGEN®) as previously described ^35,36^. PCR products were purified using the MinElute® PCR purification kit (Qiagen, United Kingdom) and sequenced with the respective forward and reverse primers in a BigDye® terminator version 3.1 (Applied Biosystems, USA) reaction and analysed in an ABI 3130xl genetic analyser.

### Sequence analysis

Raw sequence reads were quality-checked, trimmed, edited and assembled to contigs of length 420 nucleotides using Sequencher version 5.4.6 (www.genecodes.com). Alignments were prepared using MAFFT version 7.271 ^37^. IQ-TREE version 1.6.0 ^38^ was used to estimate the best fit model and infer Maximum Likelihood (ML) trees. Phylogenetic trees were generated with bootstrapping of 1000 iterations. Pairwise nucleotide p-distances were calculated using MEGA version 7.0.21 ^39^. Genotype assignment was based on the phylogenetic clustering (bootstrap value >80%) on ML trees and pairwise genetic distances to prototype strains (https://www.picornaviridae.com/enterovirus/prototypes/prototypes.htm) as proposed (10.5% for HRV-A, 9.5% for HRV-B, and 10.5% for HRV-C) ^22,23^.

Intra-type diversity for genotypes with at least ten sequences/samples was studied by visualisation of the number of nucleotide substitutions to the earliest sampled sequence for each type. The substitution rate of the VP4/2 coding region in HRV had previously been estimated as 7×10^−4^ to 4×10^−3^ substitutions/site/year ^40^. Using the upper evolutionary rate value translated to 1.68 nucleotide substitutions per year across the sequenced 420 nt segment. We therefore defined an intra-type variant as a sequence with >2 nucleotide differences from the index sequence. An intra-type variant had to be observed in at least two sequences to increase confidence that this would not be the result of sequencing error.

### Data Analysis

Data analysis was conducted using STATA version 13 (STATA Corp Texas) and R version 3.6.1 (CRAN R Project). Categorical variables were summarised using frequencies and percentages. HRV proportion for each class and respective 95% confidence intervals were defined as the number of HRV positive samples out of the total number of samples tested per class. The chi-square test for trend was used to check for linear trend in HRV proportion with an increasing hierarchy of classes in the school (from day care to teaching staff).

### Definition of terms

We defined ‘persistence’ as the continued occurrence of the same genotype within the same school term. Detection of a genotype in a subsequent school term was considered a genotype recurrence. We defined ‘frequent’ genotypes as those that occurred in at least five samples, from more than two individuals, and further investigated their temporal occurrence and persistence. We defined ‘individual HRV re-infection’ as the acquisition of a new genotype or the detection of a previously acquired genotype in a subsequent school term. Individual detection with the same genotype in consecutive samples was considered a continuing infection.

### Sequence data availability

Sequences generated by this study are available in GenBank under accession numbers MT177659-MT177911.

## RESULTS

### Baseline characteristics and HRV detection

The total number of samples collected between May 2017 and April 2018 was 1859, of which 307 (16.5%) tested positive for HRV. These HRV positives were collectively from 164 individuals: 160 pupils and four teachers. The mean age of the HRV positive pupils was 9.4 years, with a standard deviation of +3.9. The most common ARI symptom recorded among the HRV positive cases (single sampling events) was nasal discharge (n=278, 90.6%), cough (n=226, 73.6%) and sore throat (n=62, 20.2%). Only a small proportion of those detected with HRV could identify a household member with ARI-like symptoms (n=43, 14.0%). Among the household members identified, 26/43 (60%) were school-going siblings, with 18 (69%) of them attending the same school and the other eight (31%), a different school, **Table 1**. HRV circulation was detected throughout the three school terms under observation. Seasonal variations of HRV infections could not be identified due to breaks in sample collection during the school holidays. The lower primary had a higher HRV proportion compared to upper primary (19.8% versus 12.2%), *p*-value <0. 001, **Figure 1**.

**Table 1.**
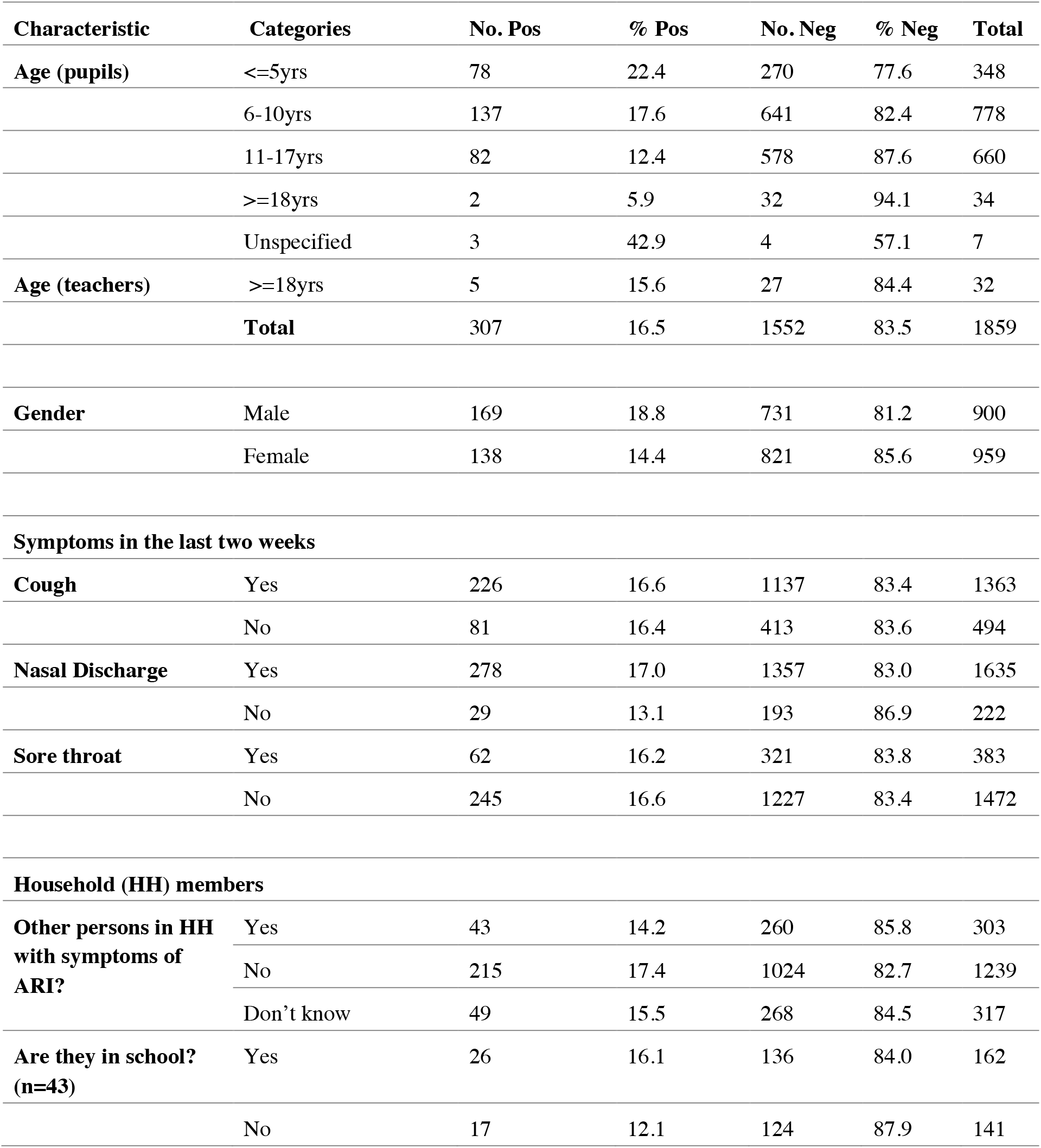
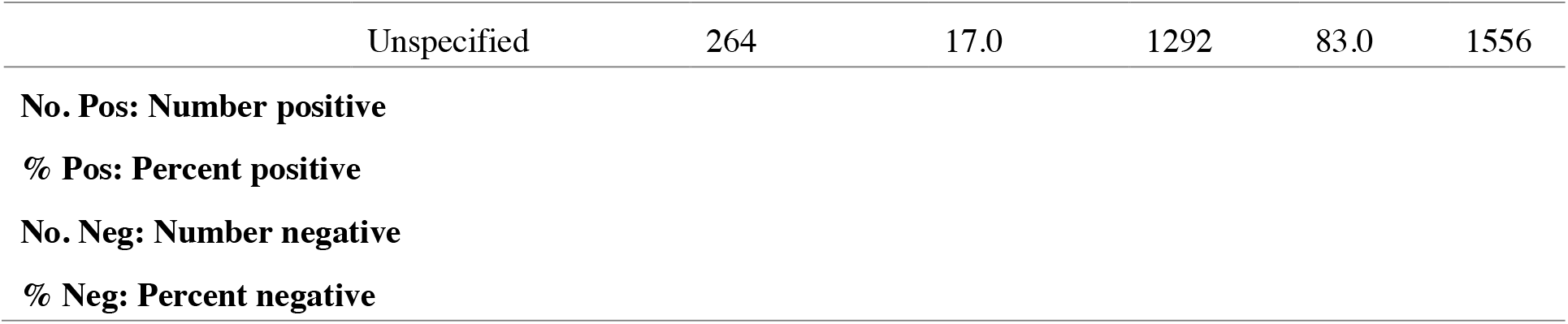
Baseline characteristics of the HRV positive cases at a rural Kenyan school sampled throughout 3 terms from May 2017 to April 2018.

**Figure 1.**
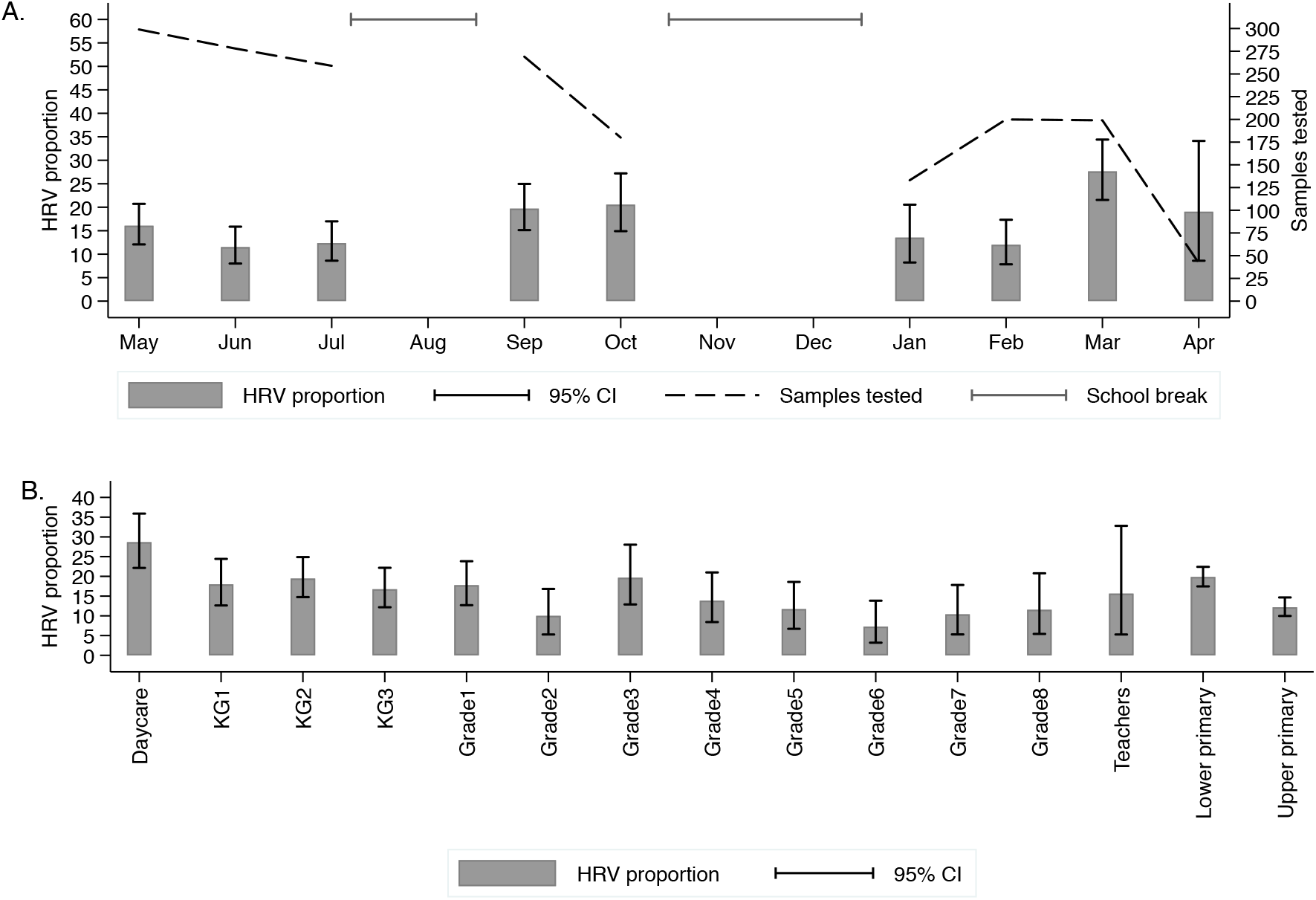
Patterns of HRV infections within the school over one year. (A). Month by month HRV proportion with respective 95% confidence intervals and number of samples tested. (B). Class-specific HRV proportion with respective 95% confidence intervals. The two bars furthest to the right are aggregated proportions of lower primary and upper primary groups.

### HRV diversity

Amplification and sequencing of the VP4/2 genomic region were attempted on all HRV positive samples resulting in 82.4% (253/307) success. The unsuccessful samples either failed to amplify or had poor sequence quality. The resulting sequences were classified into 47 HRV genotypes: 24 HRV-A genotypes, seven HRV-Bs and 16 HRV-Cs. HRV-A was the most common species (134/253, 53.0%), followed by HRV-C (73/253, 28.9%) and HRV-B (46/253, 18.2%). Some sequences violated the previously proposed genotype assignment thresholds, **Supplementary Table 1**. These were included in the analysis and classified with a suffix “-like” to the most similar known genotype (e.g. B48-like). The most frequent genotypes were A2 (24/253, 9.5%), B70 (22/253, 8.7%), A36 (16/253, 6.3%) and B48-like (16/253, 6.3%), **Figure 2**.

**Figure 2.**
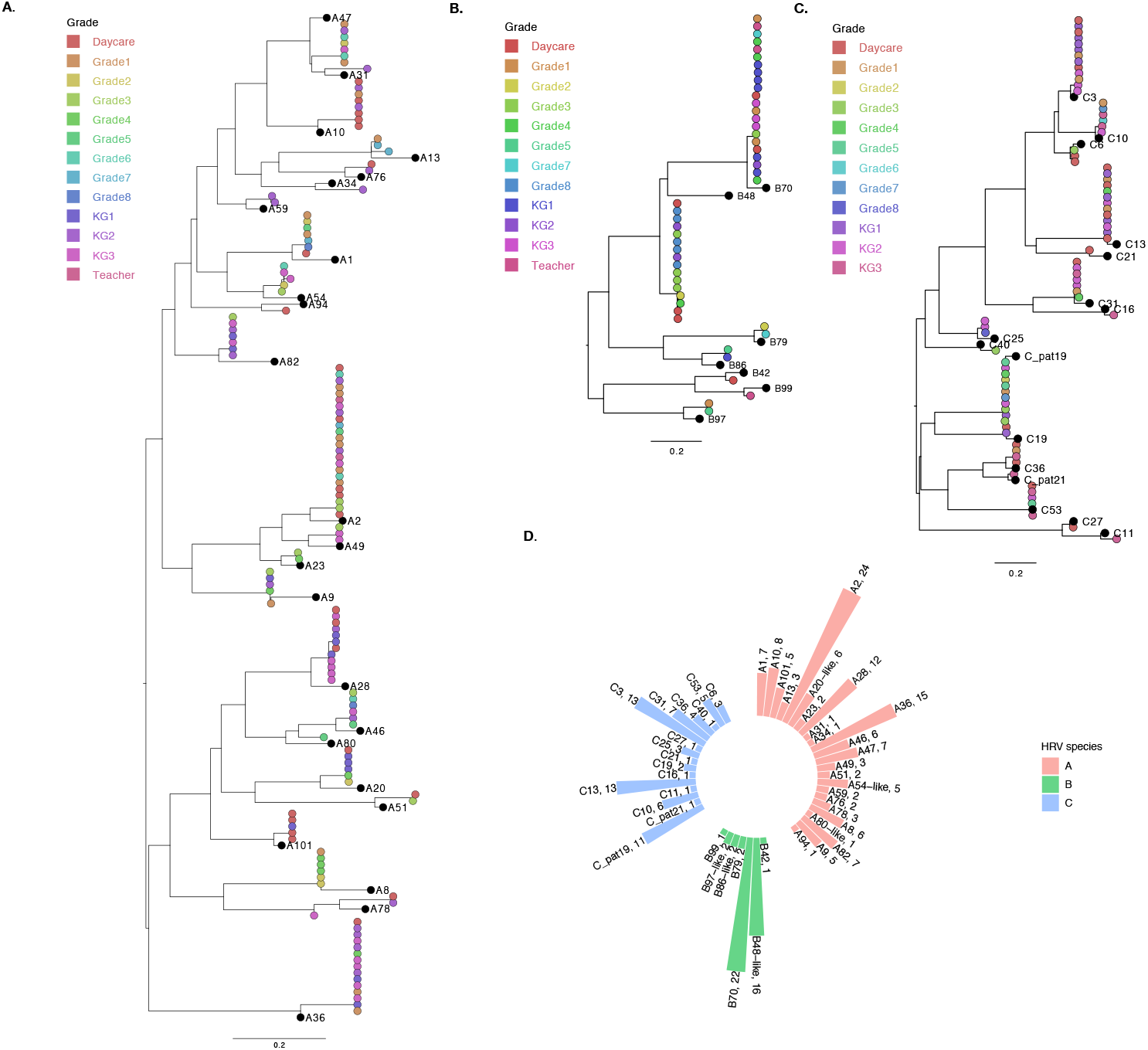
Phylogenetic analysis and genotype assignment of generated HRV sequences. Species-specific maximum likelihood trees of (A) HRV-A, (B) HRV-B and (C) HRV-C. The tip shapes are colored by the school class of the individual. The black tips represent the prototype sequence of respective strain. (D) A circular bar plot showing the frequencies of HRV genotypes identified. The bars are colored by HRV species and the tips are labeled by: HRV-type, frequency.

### Temporal occurrence and clustering patterns

Numerous genotypes circulated simultaneously in the school with 22, 15 and 19 unique genotypes observed in term one, term two and term three, respectively. Nine genotypes recurred during the study period. Of the 22 that occurred in term one, two re-occurred in term two and four in term three; whereas of the 15 observed in term two, three recurred in term three. No genotype was observed across all the three school terms. Four of the recurring genotypes (A13, A59, B48, C3) were detected in the same class, **Figure 3A**.

**Figure 3.**
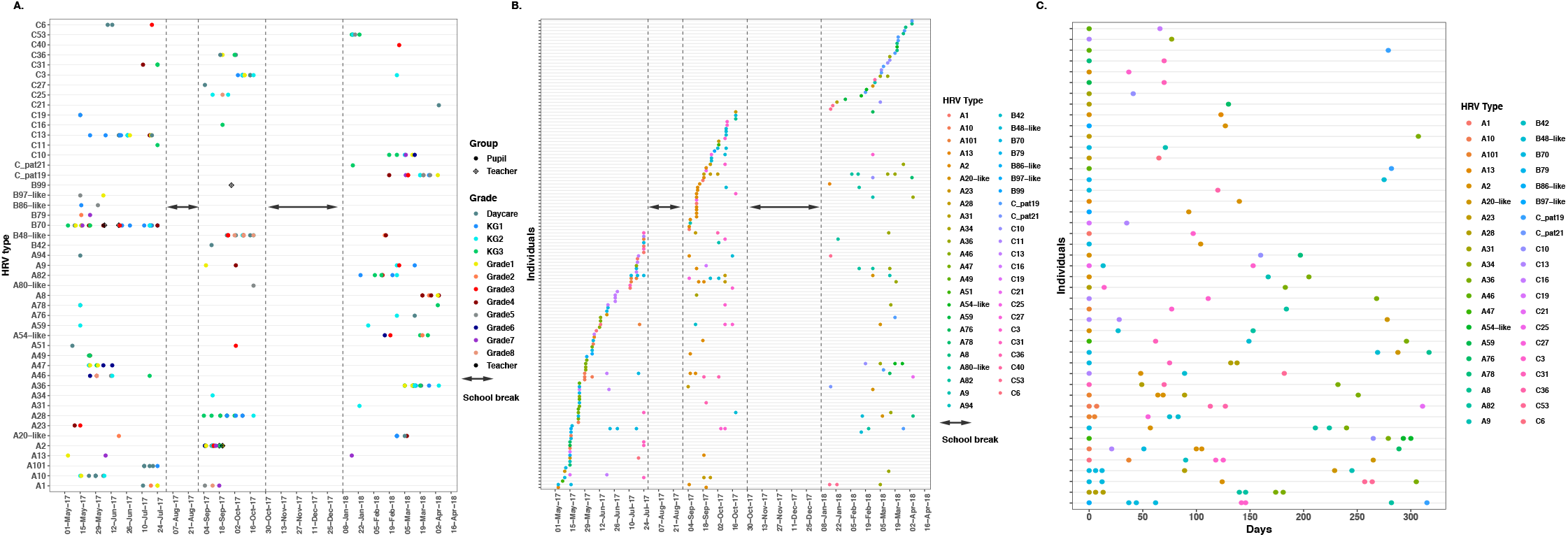
The temporal occurrence and infection patterns of HRV. (A) The temporal occurrence of all HRV genotypes across the one-year study period. (B) Individual infection patterns across the one-year period. The individuals are ordered by date of first HRV infection. (C) Number of days to a subsequent HRV case for individuals who were re-infected during the study period. All individual first infections are dated day 0. The individuals are ordered by number of HRV positive samples, with the individual with most positive samples appearing at the bottom.

Twelve genotypes were observed as singletons (i.e. present in a single sample/individual). We observed that frequent genotypes (n=21) circulated averagely for 28 days (median 23 days). Five of the frequent genotypes recurred in a subsequent school term. The longest persisting genotype was B70 (n=22 samples) seen in 81 days. Genotype A2 (n=24 samples) was similarly frequent but persisted for only 16 days. Among the frequent genotypes, none was limited to the upper primary group, whereas A10, A28, A101 and C3 were observed only in the lower primary group. However, no frequent genotype was limited to only one school class, **Error! Reference source not found**..

### Individual infection patterns

Of the 164 HRV positive individuals, 62 (37.8%) contributed more than one positive sample. Three pupils, all from the lower primary group, presented with the most HRV positive samples (n=8) per person. The 253 successfully sequenced samples were collectively contributed by 144 individuals. Repeat HRV detections (n=109) were a combination of persistent infections (24/109) and re-infections (85/109). The number of genotypes per person ranged from one to five. About two-thirds of the individuals (98/144, 68.1%) had only one HRV genotype across the study period. Overall, the highest HRV diversity per person (5 genotypes) was observed in three pupils, all from the lower primary classes, **Table 2**. Time to re-infection varied greatly (13-307 days), with a median of 77 days. However, no individual was re-infected with the same genotype across the study period **Figure 3B**.

**Table 2.**
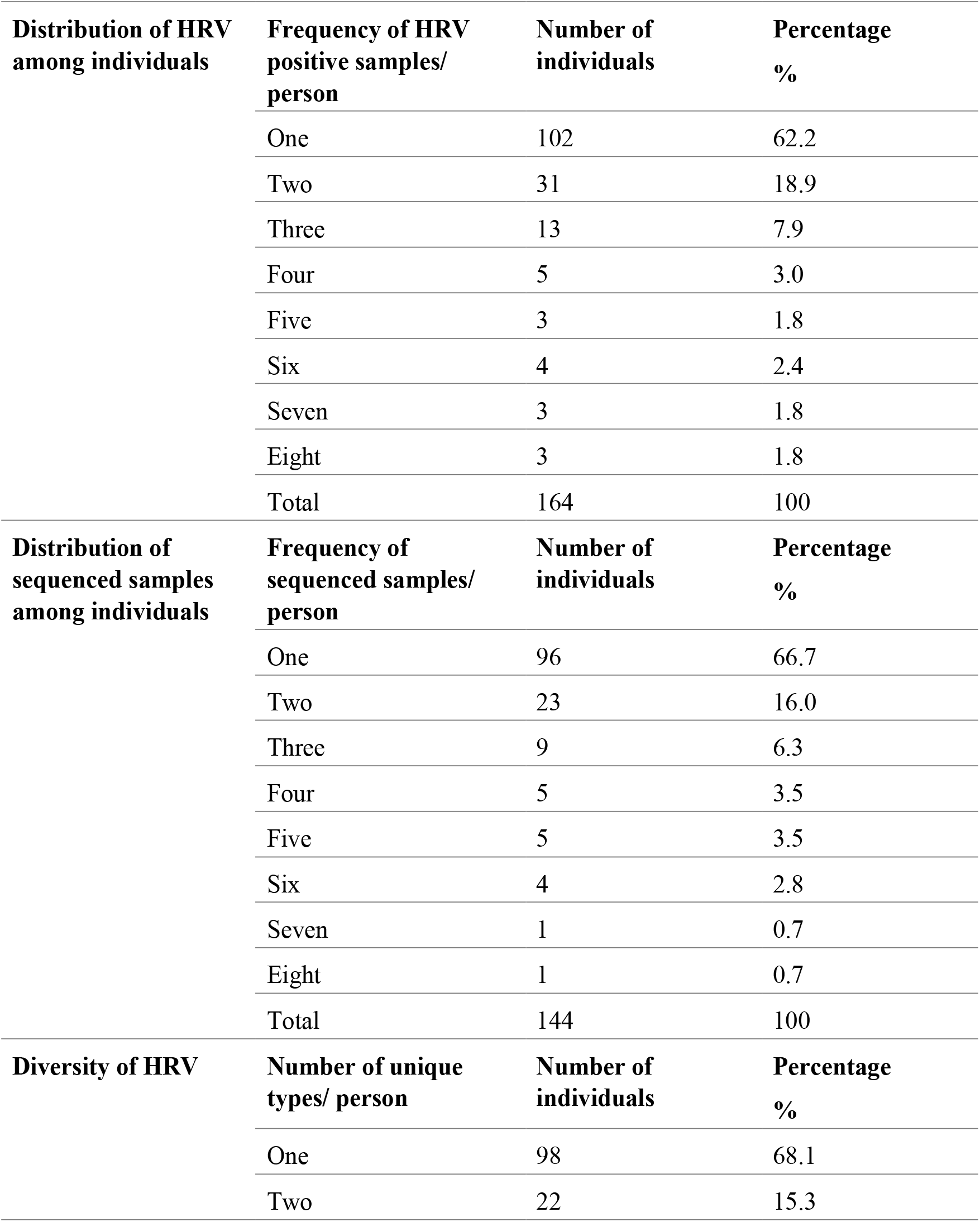
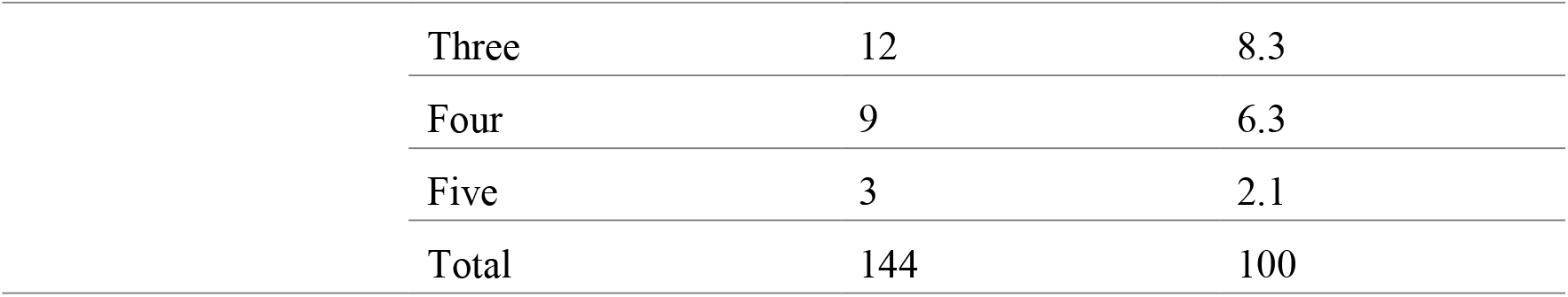
Human rhinovirus detection and genotyping summary among individuals.

| **Distribution of HRV among individuals** | **Frequency of HRV positive samples/ person** | **Number of individuals** | **Percentage %** |
| --- | --- | --- | --- |
|  | One | 102 | 62.2 |
|  | Two | 31 | 18.9 |
|  | Three | 13 | 7.9 |
|  | Four | 5 | 3.0 |
|  | Five | 3 | 1.8 |
|  | Six | 4 | 2.4 |
|  | Seven | 3 | 1.8 |
|  | Eight | 3 | 1.8 |
|  | Total | 164 | 100 |
| **Distribution of sequenced samples among individuals** | **Frequency of sequenced samples/ person** | **Number of individuals** | **Percentage %** |
|  | One | 96 | 66.7 |
|  | Two | 23 | 16.0 |
|  | Three | 9 | 6.3 |
|  | Four | 5 | 3.5 |
|  | Five | 5 | 3.5 |
|  | Six | 4 | 2.8 |
|  | Seven | 1 | 0.7 |
|  | Eight | 1 | 0.7 |
|  | Total | 144 | 100 |
| **Diversity of HRV** | **Number of unique types/ person** | **Number of individuals** | **Percentage %** |
|  | One | 98 | 68.1 |
|  | Two | 22 | 15.3 |

|  |  |  |
| --- | --- | --- |
| Three | 12 | 8.3 |
| Four | 9 | 6.3 |
| Five | 3 | 2.1 |
| Total | 144 | 100 |

### Intra-type genetic diversity

Eight genotypes had at least ten samples/sequences, and five of these occurred as a single variant throughout the study. The remaining three, A28, B48 and B70, had more than one variant. Genotype A28 had two variants simultaneously observed within the same school term, and both variants were detected only in the lower primary group. For B48, the second variant was observed as a genotype recurrence in a subsequent school term. Genotype B70 had the highest diversity, with three variants observed within one school term. The first B70 variant (13 samples) occurred across lower and upper primary as well as teaching staff, the second (4 samples) was first observed 49 days after the genotype’s overall index sequence (all samples from KG class 1) and the third (5 samples) was first observed 69 days after the genotype index sequence, with 4/5 samples coming from the lower primary. No individual had more than one variant of the same genotype, **Figure 5**.

**Figure 4.**
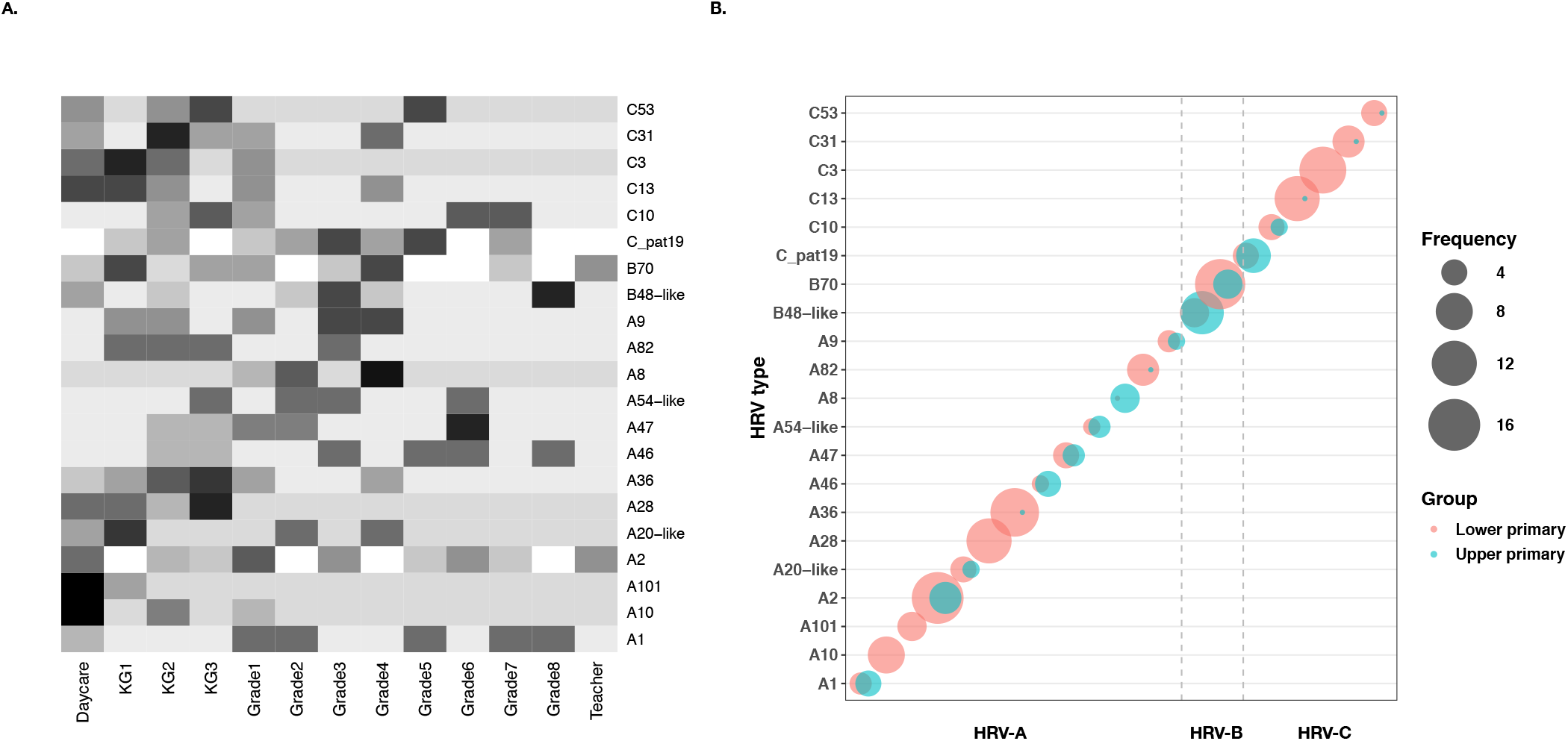
Distribution of genotypes across the school. (A) Heatmap showing the distribution of frequent genotypes across the school classes. The intensity of the color correlates to frequency of samples. Color intensity has been scaled to correct for sampling bias between the lower primary and upper primary classes. (B) Distribution of frequent genotypes between the upper and lower primary groups. The sizes of the circles correlate to number of samples and the color to the respective group: either lower or upper primary.

**Figure 5.**
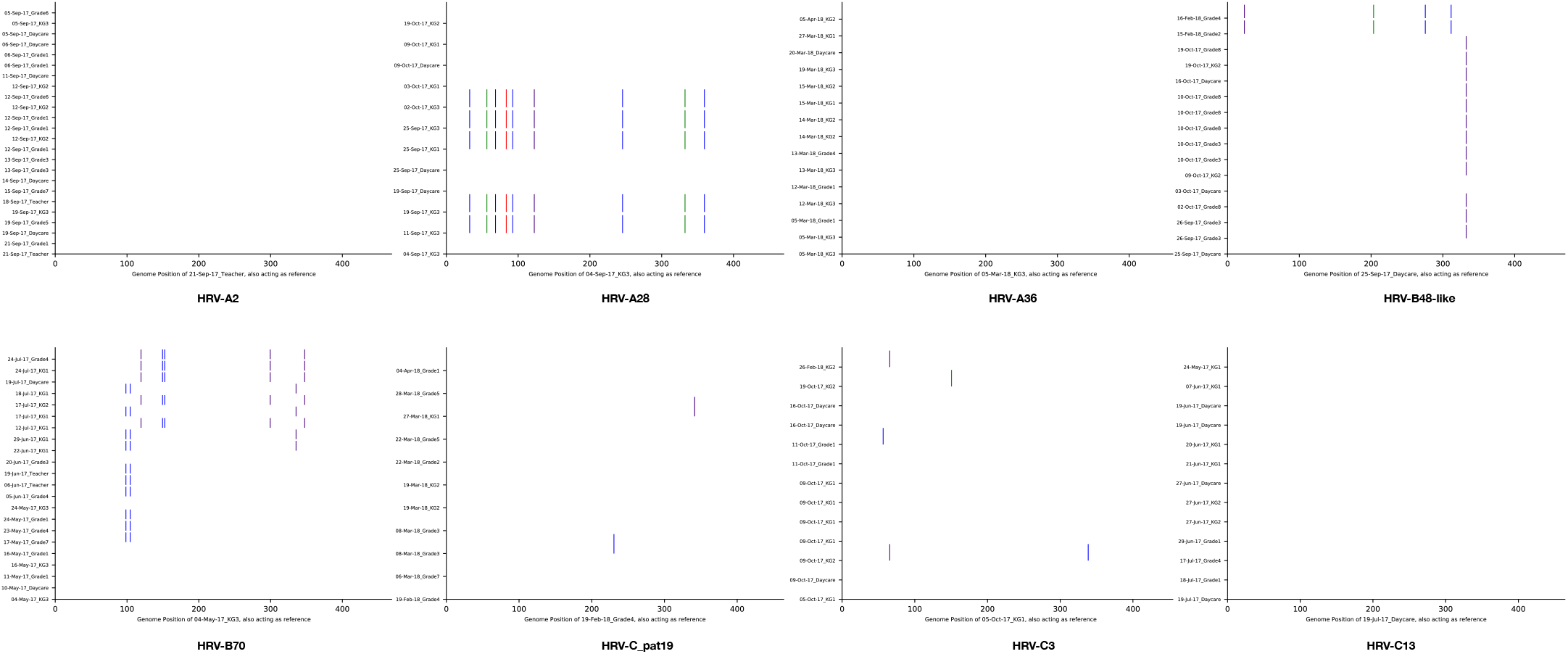
Genetic diversity of HRV genotypes with a frequency of more than ten. From top left A2, A28, A36, B48-like, B70, C_pat19, C3 and C13 (bottom right). Nucleotide substitutions are demonstrated by a colored bar. A substitution to an “A” is indicated by green, “T” by red, “G” by indigo and “C” by blue bars. The sequences are ordered by date of sample collection with the genotype’s index sequence at the bottom, and acting as a reference.

### HRV among teaching staff

Thirty-two samples were collected from teachers, of which 5 (15.6%) were HRV positive from 4 individuals. Three HRV genotypes were identified: A2 (n=2), B70 (n=2) and B99 (n=1). The A2 and B70 genotypes were detected in teachers several days after their initial detection in the student population. The B99 sample from a teacher was the only case of this genotype identified during the entire study period suggesting that they acquired the infection from outside the school setting, and no onward transmission was observed.

## DISCUSSION

This study describes the molecular epidemiology of HRV in a public primary school setting in rural coastal Kenya. We found that HRV occurs year-round in line with HRV prevalence reported among symptomatic individuals within the KHDSS (11%-23%) ^35,36,41^. HRV was detected across all age-groups, with the highest proportion in the <5 year-olds and the lowest proportion in older age groups(>=18 years), in agreement with previous findings ^25,42,43^. A proportion of the HRV positive children (14%) identified another household member as having ARI-like symptoms, suggesting transmission at the household level that might contribute to transmission at school (or vice versa).

All HRV species were found in circulation throughout the year. HRV-A circulated widely (53%), more than -B (18%) and -C (29%), a contrast to previous reports where HRV-A and HRV-C co-dominated ^35,36,41^. However, a similar occurrence of HRV species was reported in the first two years of aboriginal and non-aboriginal Australian children ^44^. There was considerable HRV diversity, with almost a third of all known HRV genotypes detected. HRV infections comprised of single genotype occurrences, observed in a single sample/individual, as well as frequent genotypes affecting numerous pupils across several classes. Previous experimental studies have shown that different HRV types possess varying degrees of infectivity ^45^.

Numerous genotypes co-circulated in every school term, implying that no particular genotype predominates at any given time period. Previous studies have shown that contemporary HRV infections in a given population are characterised by numerous genotype-specific “mini-epidemics”^46^ and that up to 30 genotypes circulate simultaneously at a given geographical area^47^. No frequent genotype was limited to one class, suggesting heterogeneous mixing and transmission within the school. However, four frequent genotypes and one variant of B70 were observed only in the lower primary, an indication of social clustering. Genotype recurrence in a subsequent school term was observed in nine genotypes. Although it is not clear whether the study design missed samples between two genotype occurrences, the infrequency of genotype recurrence is possibly a reflection of herd immunity to specific types within the school/local community or a reflection of random introductions into the school/local community. Frequent genotypes in the school persisted for about a month on average. This is a shorter period than earlier observed across the KHDSS (a larger geographical scope) during an outpatient surveillance ^41^. This is probably due to increased transmission (steered by high contact rates among school-going children) resulting in a shorter duration epidemic.

The younger age groups exhibited high rhinovirus diversity as they had more HRV re-infections. No individual was re-infected with the same genotype; further evidence of serotype-specific immunity to HRV lasting at least one year ^48^.

We demonstrate the occurrence of intra-genotype variants, which were either separate rhinovirus introductions or diversification of a single variant after introduction and as a result forming different transmission clusters. This observation highlights the benefit of sequence data over serology to study viral transmission dynamics.

An outpatient health facility located within the same location as the school was recruited into an ARI surveillance study ^49^ from December 2015 to November 2016, five months prior to our school study. This outpatient clinic is within a radius of 4 km from the school. Detailed analysis of molecular epidemiology of HRV for samples collected at this outpatient clinic was reported elsewhere ^41^. Although not a primary objective of this study, we compared the diversity of HRV infections between the two study periods. We observed 12 common genotypes in the two studies: A13, A20, A28, A31, A46, A54, A78, A101, B42, C6, C11 and C19. However, only one genotype was frequent in both periods: A101 (**Supplementary figure 1**). Our comparison of HRV diversity between a school setting and clinical cases in a health facility within the same geographical location and in two consecutive seasons showed only one frequent genotype present in both studies. This is an indication that HRV diversity within a community varies widely over time, as previously observed ^50^. It is not definite what drives the exchange of common rhinovirus genotypes. The rapid turnover and co-existence of genotypes and variants might be determined by immunologically mediated selection processes or other nonselective epidemiological processes.

Our study had some limitations. First, the dichotomy in the number of samples collected weekly in lower primary versus upper primary posed a challenge when comparing the two groups. Second, weekly sampling of only symptomatic persons will likely have resulted in missed HRV infections, impairing the overview of HRV dynamics. In addition, the study failed to successfully amplify and sequence nearly 18% of HRV positive samples. Failure was not correlated with viral load and may have been caused by variability in primer-annealing sites resulting in mismatches. This may have resulted in missed genotypes or sub-variants.

This study provides improved knowledge of the diversity and temporal characteristics of HRV in a school setting, reinforcing the notion that schools are a focal point in understanding HRV transmission in the community. The effect of numerous individuals in close contact enabling HRV transmission is evident. In addition, we see that infections could be linked to transmission events occurring outside the school setting, i.e. household setting. The contemporary inclusion of different population structures (e.g. schools, households, health centres) in studying HRV dynamics will improve understanding of HRV epidemiology in communities. Future studies should focus on whole-genome sequencing to fully elucidate transmission clusters.

## Data Availability

The replication data and analysis scripts for this manuscript shall be made available at the Harvard Dataverse: (https://dataverse.harvard.edu/dataverse/vec). Some of the clinical dataset contains potentially identifying information on participants and is stored under restricted access. Requests for access to the restricted dataset should be made to the Data Governance Committee of the KEMRI-Wellcome Trust Research Programme.

## Data availability

The replication data and analysis scripts for this manuscript shall be made available from the Harvard Dataverse: (https://dataverse.harvard.edu/dataverse/vec). Some of the clinical dataset contains potentially identifying information on participants and is stored under restricted access. Requests for access to the restricted dataset should be made to the Data Governance Committee.

## Acknowledgements

We thank all the study participants for their contribution of samples and data. We also thank the School teachers, School Management Board, Parent Teacher Association and County Education Management Committees for allowing us to conduct the study within their school. We are grateful to the field study team for participant recruitment and the laboratory staff of the KEMRI-Wellcome Trust Research Programme / Virus Epidemiology and Control research group. This paper is published with the permission of the Director of KEMRI.

## Supplementary Material

**Supplementary Table 1:** Limits of intra-type pairwise genetic distances of Kilifi school sequences to HRV prototype strains

| Type | Accession number of prototype strain | Number of sequences | Upper limit p-distance |
| --- | --- | --- | --- |
| A1 | D002390.1 | 7 | 0.093 |
| A10 | DQ4734980.1 | 8 | 0.071 |
| A101 | GQ4150510.1 | 5 | 0.044 |
| A13 | FJ4451160.1 | 3 | 0.099 |
| A2 | X023160.1 | 24 | 0.102 |
| A20-like * | FJ4451200.1 | 6 | 0.118 |
| A23 | DQ4734970.1 | 2 | 0.074 |
| A28 | DQ4735080.1 | 12 | 0.102 |
| A31 | FJ4451260.1 | 1 | 0.104 |
| A34 | DQ4735010.1 | 1 | 0.093 |
| A36 | DQ4735050.1 | 15 | 0.091 |
| A46 | DQ4735060.1 | 6 | 0.071 |
| A47 | FJ4451330.1 | 7 | 0.085 |
| A49 | DQ4734960.1 | 3 | 0.102 |
| A51 | FJ4451360.1 | 2 | 0.088 |
| A54-like * | FJ4451380.1 | 5 | 0.107 |
| A59 | DQ4735000.1 | 2 | 0.088 |
| A76 | DQ4735020.1 | 2 | 0.082 |
| A78 | EF1734180.1 | 3 | 0.102 |
| A8 | FJ4451130.1 | 6 | 0.085 |
| A80-like* | FJ4451560.1 | 1 | 0.137 |
| A82 * | DQ4735090.1 | 7 | 0.113 |
| A9 | FJ4451770.1 | 5 | 0.077 |
| A94 | EF1734190.1 | 1 | 0.102 |
| B42 | FJ4451300.1 | 1 | 0.073 |
| B48-like * | DQ4734880.1 | 16 | 0.154 |
| B70 | DQ4734890.1 | 22 | 0.09 |
| B79 | FJ4451550.1 | 2 | 0.051 |
| B86-like * | FJ4451640.1 | 2 | 0.121 |
| B97-like * | FJ4451720.1 | 2 | 0.118 |
| B99 | FJ4451740.1 | 1 | 0.076 |
| C pat19 | FJ5980960.1 | 11 | 0.047 |
| C pat21 | FJ6157370.1 | 1 | 0.061 |
| C10 | GQ3237740.1 | 6 | 0.05 |
| C11 | EU8409520.1 | 1 | 0.087 |
| C13 | EU0817950.1 | 13 | 0.041 |
| C16 | EU0818080.1 | 1 | 0.047 |
| C19 | EU6978500.1 | 2 | 0.055 |
| C21 | EU7523770.1 | 1 | 0.073 |
| C25 | EU7524270.1 | 3 | 0.105 |
| C27 | GQ2231220.1 | 1 | 0.026 |
| C3 | EF1860770.1 | 13 | 0.052 |
| C31 | GU2943800.1 | 7 | 0.079 |
| C36 | EF0772560.1 | 4 | 0.093 |
| C40 | EU0818020.1 | 1 | 0.064 |
| C53 | MF7753670.1 | 5 | 0.006 |
| C6 | EF5823870.1 | 3 | 0.061 |
| * Do not conform to proposed VP4/2 thresholds. This has been documented for genotypes A78 (intra-clade upper limit of 0.153), A82 (intra-clade upper limit of 0.135) and A20 (intra-clade upper limit of 0.119) <sup>23</sup>. |  |  |  |

## Supplementary Figure legend

**Supplementary Figure 1:**
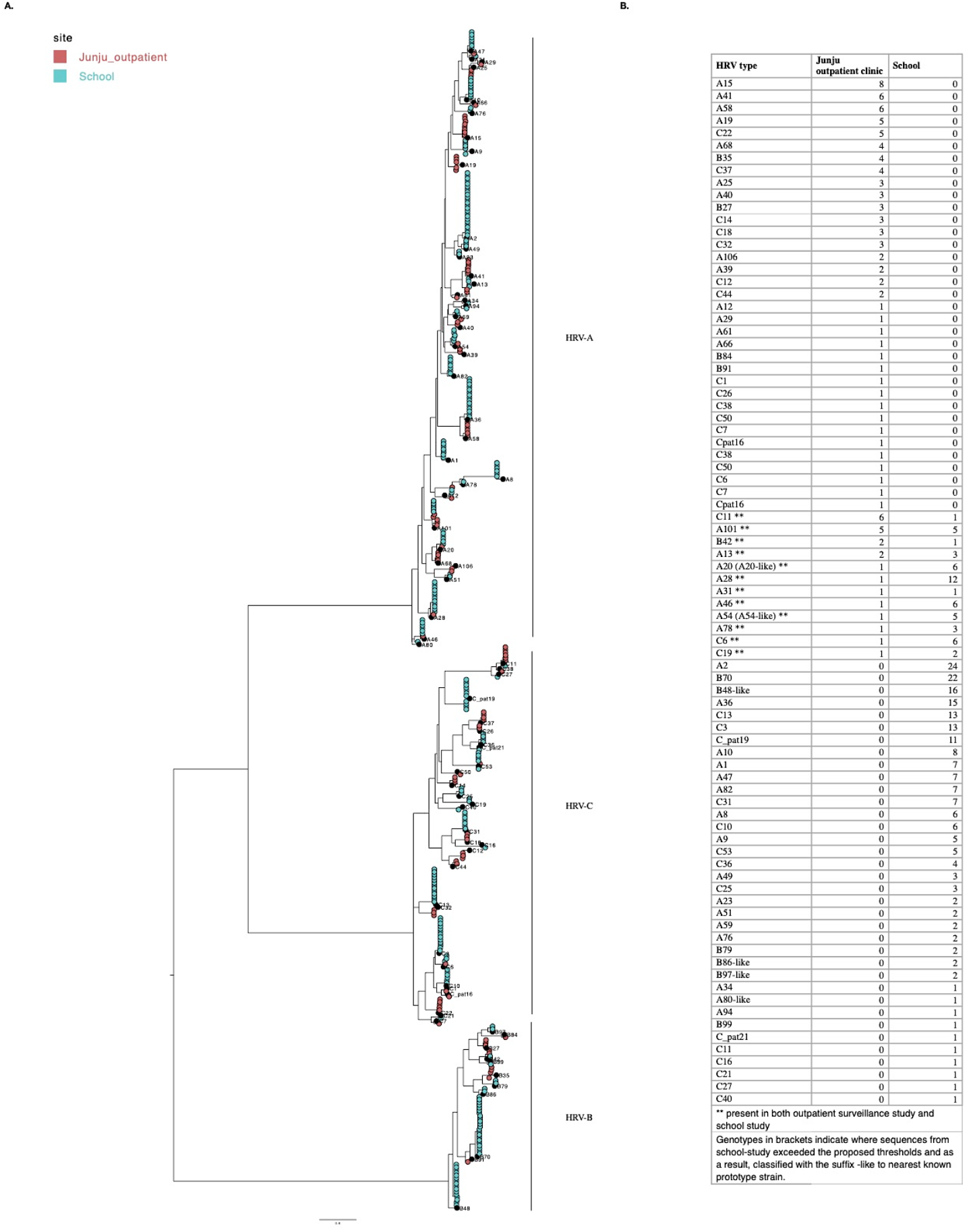
Genotype distribution between the school and the Junju outpatient clinic. (A). Phylogenetic analysis of sequences from the two studies. The tips are colored by study site. (B). A table with genotype frequencies from both studies. Only 12 genotypes were present in both studies.

